# Timing of antidepressant use influences long term functional status in New Zealand stroke patients: A retrospective population level study

**DOI:** 10.1101/2025.07.25.25332233

**Authors:** Shivankar Nair, Emma M. Tuckey, Deepak Gupta, Rong Liu, Alesha J. Smith, Ailsa L. McGregor

**Affiliations:** School of Pharmacy, University of Otago, Dunedin, New Zealand; Chengdu University of Traditional Chinese Medicine, Chengdu, China

**Keywords:** Stroke, antidepressant, functional status, venlafaxine, SSRI

## Abstract

This retrospective analysis explored the relationship between the timing of antidepressant use and long-term functional status after stroke.

We used linked health administrative data from a cohort of adult stroke patients in New Zealand. Demographics and prescription information were obtained from the National Minimum Dataset and Pharmaceutical Collection. Activities of Daily Living (ADL) scores for the same patients were obtained from the International Resident Assessment Instrument (InterRAI™). Beta regression investigated any relationship between antidepressant exposure and functional status.

Of 3509 patients with an ischaemic stroke, 31% used antidepressants in the three months before or after stroke. The adjusted odds ratio (OR) for exposure before and after stroke was 0.92 (95% confidence interval [CI]: 0.83-1.01) and 1.19 (95% CI: 1.06-1.31) for post-stroke exposure. Tricyclic antidepressant (TCA) or venlafaxine use after stroke was associated with greater odds of a lower ADL score compared to selective serotonin reuptake inhibitors (SSRI).

Patients prescribed antidepressants after stroke had increased odds of higher ADL scores indicating poorer long-term functional status than those who used them before and after stroke or not at all. TCAs and venlafaxine appeared less detrimental to long term function than SSRIs and may be better options for managing post stroke depression.

**Key take-home messages:**

1. Timing of antidepressant use impacts stroke recovery – Patients who started antidepressants only after stroke had poorer long-term functional outcomes compared to those who used them before and after stroke or not at all.
2. Choice of antidepressant matters – Tricyclic antidepressants (TCAs) and venlafaxine were associated with better functional outcomes than SSRIs, suggesting that SSRIs may not be the best option for post-stroke depression management.
3. Poorer functional outcomes were not due to stroke severity – No difference in hospital length of stay between groups suggests that the negative impact of post-stroke antidepressant use on function is not simply due to more severe strokes.
4. Future research should guide prescribing decisions – More studies are needed to understand whether antidepressants directly influence recovery or if patient characteristics drive these outcomes, helping to refine treatment strategies for optimising both mental health and functional recovery in stroke survivors.

## 1. Introduction

Depression after stroke is a significant clinical issue affecting approximately 27% of patients.(1) Post-stroke depression is associated with poor prognosis and increased mortality.(2) Selective serotonin reuptake inhibitors (SSRIs) are the first-line pharmacological treatment to manage depression after stroke,(3) however, there is little evidence to support the use of a particular SSRI and the optimal timeframe for initiation and duration of treatment remains unclear.(4)

Independent of their effect on depression, SSRIs have been shown to alter the excitatory– inhibitory balance in periinfarct tissue and reinstate a form of plasticity like that observed in the critical period in development.(5) Meta-analysis studies have however failed to reach a consensus on whether exposure to SSRIs can promote recovery in stroke survivors.(6, 7)

Despite definitive evidence of benefit, there has been a growing interest in how the timing of SSRI use relative to stroke affects recovery. Prior SSRI use was associated with a lower likelihood of being discharged to home.(8) In contrast, patients with prior SSRI use have shown better short-term outcomes compared to those with a new SSRI prescription after stroke.(9) The beneficial impact of prior exposure on stroke recovery is further supported by the observation that use of any class of antidepressant medicine in the week before stroke was associated with a greater chance of returning to baseline function at 90 days.(10)

Taken together, these results suggest that exposure to antidepressant medicine influences the short-term recovery trajectory. However, the impact of antidepressant treatment on longer term functional status has not been investigated. The objective of this study was to investigate any association between the timing of antidepressant treatment and functional status assessed by ADL scores in New Zealand stroke patients.

## 2. Methods

### 2.1 Study population

Linked health administrative data formed a population-based cohort of 3509 stroke patients in New Zealand. A case definition of at least one hospital admission for ischaemic stroke (ICD-10 code I60–I69) from 1 January 2017 to 31 December 2022, regardless of age, was adopted to identify patients in the National Minimum Dataset (NMDS). Patients were removed from the dataset if they experienced more than one stroke in the study period. Variables included: age, sex, ethnicity, date of hospital admission and length of hospital stay. The first admission date was considered the stroke diagnosis date. Prescription dispensing records from the Pharmaceutical Collection were linked for each patient’s three months pre and post stroke admission date. The NMDS and Pharmaceutical Collection capture hospitalisation information and community dispensed prescription medicines for approximately 96% of New Zealanders.

Medicines in the Pharmaceutical Collection were sorted into classes based on their primary indication (Supplementary Table 1).(11) Dosage information was used to confirm the class for medicines with multiple indications.

Functional status was assessed using a standardised ADL score extracted from InterRAI™ and linked for each patient using National Health Index (NHI) numbers (a unique health identifier used for a patient’s lifetime, across all aspects of the health system). The National Ethics Advisory Committee’s Ethical Guidelines permits the re-use of de-identified Ministry of Health data for research with a written consent waiver.

### 2.2 Outcome variable

Total ADL Score (range 0-60) was the primary outcome, a summed score from ten subscales (bathing, personal hygiene, dressing upper body, dressing lower body, walking, locomotion, transfer toilet, toilet use, bed mobility and eating). Scores for each subscale ranged from zero (independent) to six (dependent) or eight (the activity did not occur). For scores of 8, total dependence was assumed.(12)

Patient with a functional assessment date only before their stroke diagnosis date were removed. Only the first assessment after the stroke diagnosis date was considered.

### 2.3 Antidepressant exposure definition

The independent variable was exposure to antidepressant medicine approved for the management of depression and anxiety in New Zealand. A ternary variable for antidepressant exposure that reflected different clinical scenarios was created using Pharmaceutical Collection data,

- ‘Before and after stroke’ - patients dispensed an antidepressant in the 90 days preceding and in the 90 days after stroke (post-depression stroke).
- ‘Only after stroke’ – patient dispensed an antidepressant ≤ 90 days after stroke (post-stroke depression)
- ‘Unexposed’ - patient not dispensed an antidepressant during this period (reference group).

Patients exposed to antidepressants only before their stroke (n=86) were removed from primary analysis.

### 2.4 Covariates

Biological variables associated with functional ability were included.(13) Age in years on stroke diagnosis date was categorised into 40-55, 56-70, 71-85 and over 85 years. Body mass index (BMI) was derived from height and weight measurements in interRAI™. BMI was imputed for patients with missing height or weight measurement. Patients with a calculated BMI outside the feasible range of 12-100 were excluded (Figure 1A). BMI was categorised as underweight (<18.50), normal weight (18.50-24.99), overweight (25.00-29.99) and obese (≥ 30.00) consistent with New Zealand thresholds.(14) Sex was categorised as male or female and ethnicity as European, Māori, Pacific People, Asian and other.

**Figure 1.**
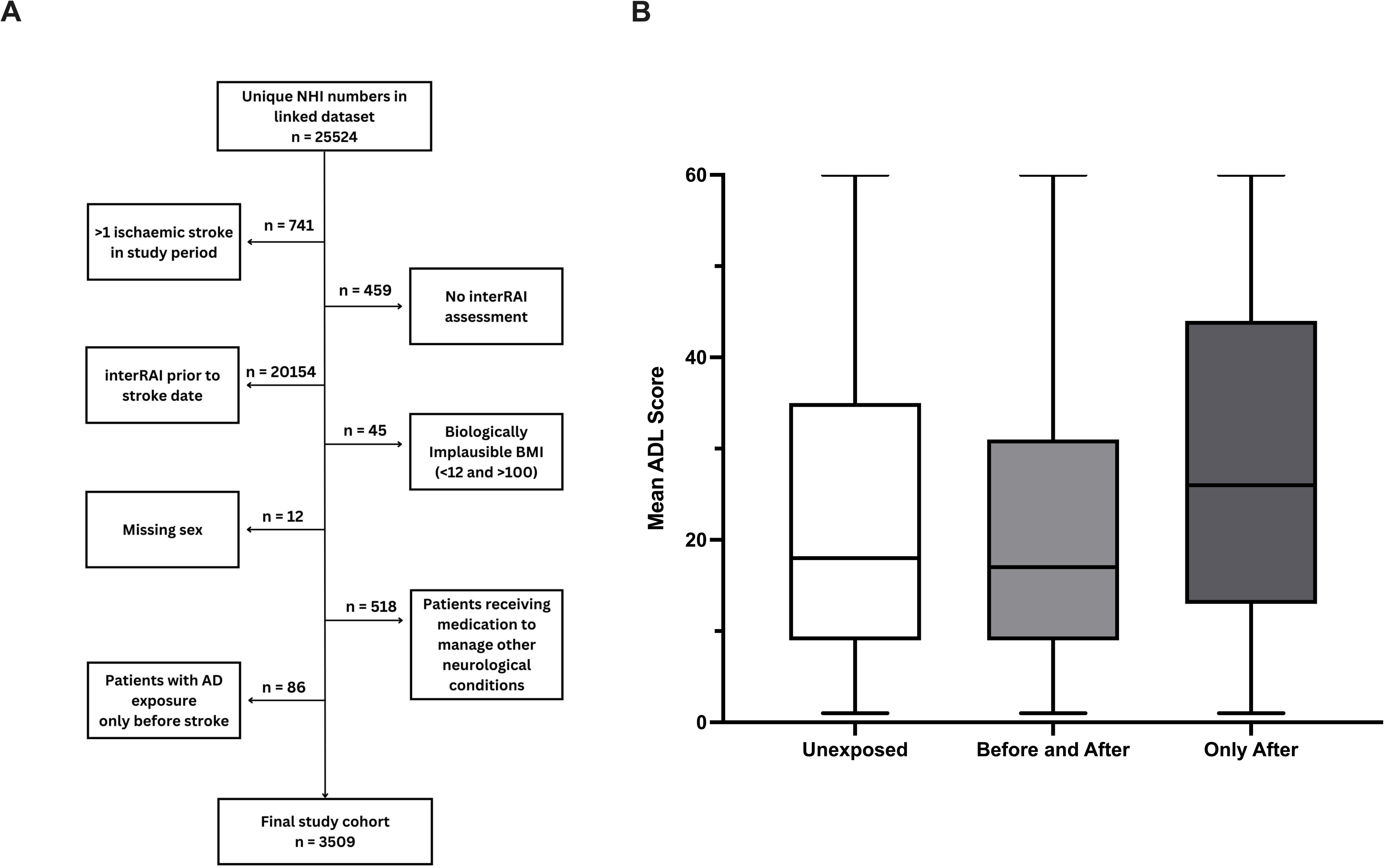
A total of 3509 patients were included in the study (**A**). The median total ADL score was higher in patients exposed to antidepressants only after stroke, and scores in the upper quartile varied less compared with other exposures. The median ADL score and interquartile range in patients exposed both before and after stroke was comparable to that of unexposed patients (**B**). AD: antidepressant, ADL: activities of daily living, BMI: body mass index, NHI: national health index, interRAI™: international residential assessment instrument

Both the severity of the initial stroke and the timing of the assessment following stroke impact ADL scores. Consequently, total length of hospital stay (TLoS, a surrogate marker for initial stroke severity, (15, 16) and time to assessment (TTA) were included.

Binary exposure to other centrally acting medicines that may affect function after stroke was included (Supplementary Table I).(17) Patients who received medicines for Parkinson’s disease, Huntington’s disease, dementia or who received aprepitant to manage post-operative nausea and nausea related to chemotherapy were also excluded.

### 2.5 Statistical analysis

Baseline characteristics for antidepressant exposed and unexposed stroke patients were compared using chi-squared tests for categorical variables or Wilcoxon’s rank-sum tests for continuous variables.

To account for the non-normal and highly skewed distribution of ADL scores, Beta regression was used to investigate the relationship between the timeframe of exposure to antidepressant medicine and functional status and exposure to specific antidepressant classes in the post-stroke timeframe (SSRIs, TCAs, mirtazapine and venlafaxine) and functional status in New Zealand stroke patients. Exposure to monoamine oxidase inhibitors (MAOI) after stroke was removed from antidepressant class analysis due to the small number of patients (n=2).

The mean models were adjusted for: sex and age category, categorical BMI, ethnicity, TLoS, TTA, and exposure to medicines that may affect post-stroke functional ability including NSAIDs, paracetamol, opioids, antiepileptics, anti-psychotics, hypnotics, anti-muscarinic, anti-nausea and smoking cessation medicines. Logit link function and log link functions were used for mean and precision model components. Variance inflation factor (VIF) and likelihood ratio (LR) test were considered to determine which covariates should be included in adjusted precision models. (18, 19)

Graphpad Prism Version 10.4.1. (SAS Institute, Cary, North Carolina, USA) and R Version 4.4.2 were used for all analyses. All effect estimates are presented with 95% confidence intervals (95% CI). Significance was assumed with a *p* value of <0.05.

## 3. Results

We identified 3509 patients who suffered an ischaemic stroke in New Zealand from January 2017 to December 2022 who fulfilled the inclusion criteria. Our total population was 54% female. Of those, 1192 (32%) were exposed to an antidepressant either after stroke or both before and after their stroke diagnosis. The remaining 2403 (68%) stroke patients did not receive an antidepressant and were thus unexposed (Table 1).

**Table 1:** Patient characteristics. Data is presented as Mean ± SD or N (%) unless otherwise stated. HC Home Care, LTCF Long Term Care Facility, TLoS Total Length of Hospital Stay, TTA
Time to Assessment

| Variable | Total sample<br>(n = 3509) | Antidepressant exposure category |  |  | *p* value |
| --- | --- | --- | --- | --- | --- |
|  |  | Unexposed<br>(n = 2403) | Before and After<br>(n = 551) | Only After<br>(n = 555) |  |
| **Age category (Years)** |  |  |  |  | **< 0.001** |
| 40-55 | 26 (0.8%) | 13 (0.5%) | 7 (1.3%) | 6 (1.1%) |  |
| 56-70 | 319 (9.1%) | 191 (7.9%) | 66 (12%) | 62 (11%) |  |
| 71-85 | 1676 (48%) | 1076 (45%) | 289 (52%) | 311 (56%) |  |
| >85 | 1488 (42%) | 1123 (47%) | 189 (34%) | 176 (32%) |  |
| **BMI Category** |  |  |  |  | **0.060** |
| Underweight | 395 (11%) | 277 (12%) | 58 (11%) | 60 (11%) |  |
| Normal | 970 (28%) | 684 (28%) | 140 (25%) | 146 (26%) |  |
| Overweight | 1567 (45%) | 1074 (45%) | 237 (43%) | 256 (46%) |  |
| Obese | 577 (16%) | 368 (15%) | 116 (21%) | 93 (17%) |  |
| **Sex** |  |  |  |  | **0.2** |
| Male | 1609 (46%) | 1123 (47%) | 235 (43%) | 251 (45%) |  |
| Female | 1900 (54%) | 1280 (53%) | 316 (57%) | 304 (55%) |  |
| **Ethnic Group** |  |  |  |  | **< 0.001** |
| Asian | 188 (5.4%) | 141 (5.9%) | 22 (4%) | 25 (4.5%) |  |
| European | 2834 (81%) | 1892 (79%) | 490 (89%) | 452 (81%) |  |
| Māori | 325 (9.3%) | 243 (10%) | 30 (5.4%) | 52 (9.4%) |  |
| Pacific peoples | 162 (4.6%) | 127 (5.3%) | 9 (1.6%) | 26 (4.7%) |  |
| **Number of Medications Taken** |  |  |  |  | **< 0.001** |
| 1-4 | 2812 (80%) | 2127 (89%) | 308 (56%) | 377 (68%) |  |
| 5-9 | 667 (19%) | 270 (11%) | 228 (41%) | 169 (30%) |  |
| 10+ | 30 (0.9%) | 6 (0.2%) | 15 (2.7%) | 9 (1.6%) |  |
| **Location of Assessment** |  |  |  |  | **0.044** |
| HC | 3245 (92) | 2234 (93.0) | 512 (93) | 499 (90) |  |
| LTCF | 264 (7.5) | 169 (7.0) | 39 (7.1) | 56 (10) |  |
| **TLoS (days)** | 8 ± 8 | 8 ± 8 | 8 ± 7 | 9 ± 8 | **0.035** |
| **TTA (days)** | 200 ± 285 | 205 ± 288 | 225 ± 291 | 155 ± 261 | **<0.001** |

Data is presented as Mean ± SD or N (%) unless otherwise stated.

### 3.1 Background characteristics

Patients in the antidepressant-exposed groups were generally younger than unexposed patients; around half of the patients in the unexposed group were aged over 85 years (47%), compared to approximately one third of patients exposed before and after (34%), or only after (32%).

While the majority of patients in all exposure groups identified as European (81%), those exposed to an antidepressant both before and after stroke were less likely to be Māori (5.4% [before and after] vs 9.4% [only after] and 10% [unexposed]) or Pacific peoples (1.6% [before and after] vs 4.6% [only after] and 4.7% [unexposed]).

A higher percentage of patients exposed to antidepressants took five or more other centrally acting medicines compared to unexposed patients (43.7% [before and after] and 31.6% [only after] vs 11.2% [unexposed]).

TLoS was comparable between the three exposure categories, however, the mean time to assessment was shorter for patients exposed only after stroke (155 ± 261 days) compared to those exposed both before and after (225 ± 291 days) and unexposed patients (205 ± 288 days, p<0.001). Patients exposed only after stroke were also more likely to be assessed in a long-term care facility (10% vs 7.1% [before and after] and 7% [unexposed]).

### 3.2 The effect of antidepressant timing on the functional status of stroke patients

Patients exposed to antidepressants only after stroke had a median ADL score of 26 (IQR 13-44) while median ADL scores were comparable for unexposed patients and those exposed to antidepressants both before and after stroke (18 [IQR 9-35] and 17 [IQR 9-31) respectively, Figure 1B).

Beta regression showed that exposure to antidepressants only after stroke was significantly associated with greater odds of having a higher ADL score than non-exposure: unadjusted odds ratio (OR) 1.30 (95% CI 1.18–1.44). After adjusting for covariates, the OR was 1.19 (95% CI 1.06–1.31). Further, the unadjusted and adjusted OR for antidepressant exposure both before and after stroke was 0.92 (95% CI 0.83–1.02) and 0.92 (95% CI 0.83–1.01), respectively (Table 2).

**Table 2:** Beta regression – Antidepressant exposure by timeframe. Figures in parentheses are 95% confidence intervals. OR: odds ratio. The mean and precision components of the model were adjusted for age, sex, ethnicity, body mass index, length of hospital stay, and other medicines. All centrally acting medicines were considered in the mean model; the precision model used pain and antipsychotic medicines.

| Exposure | Unadjusted effect |  | Adjusted effect |  |
| --- | --- | --- | --- | --- |
|  | OR Estimate | p-value | OR Estimate | p-value |
| **Mean model (n = 3509)** |  |  |  |  |
| Unexposed | Reference |  | Reference |  |
| After only | 1.30 (1.18-1.44) | **<0.001** | 1.19 (1.06-1.31) | **<0.001** |
| Both before and after | 0.92 (0.83-1.02) | 0.118 | 0.92 (0.83-1.01) | 0.078 |
| **Precision model (n = 3509)** |  |  |  |  |
| Unexposed | Reference |  | Reference |  |
| After only | 0.04 (-0.07-0.15) | 0.450 | 0.04 (-0.07-0.15) | 0.471 |
| Both before and after | 0.09 (-0.02-0.21) | 0.102 | 0.10 (-0.02-0.22) | 0.089 |

### 3.3 Frequency of use of antidepressant classes in exposed patients

The frequency distributions of different medicine classes in antidepressant exposed individuals are shown in Figure 2.

**Figure 2.**
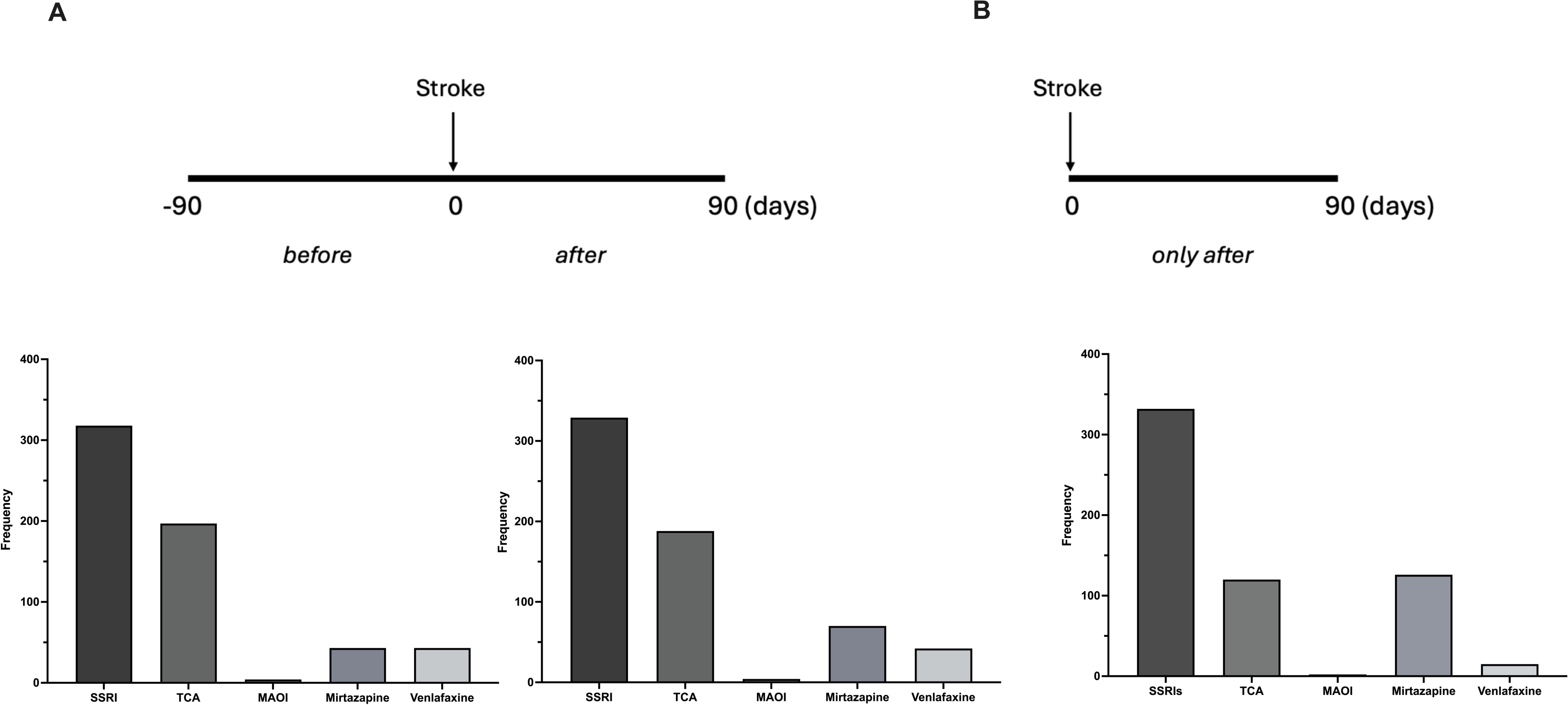
Visual timeline of antidepressant exposure and frequency of prescriptions in each antidepressant class before and after stroke (A) and only after stroke (B). After stroke, prescribing patterns remained similar, but mirtazapine prescriptions increased, while TCA and venlafaxine prescriptions decreased for patients exposed only after stroke.

The pattern of antidepressant prescribing before and after stroke was similar (Figure 2A), with SSRIs most commonly prescribed. MAOI prescribing remained minimal. Post-stroke, there were occasional medicine changes. Most common were switches from TCAs to SSRIs and between different SSRIs. Mirtazapine was frequently added for patients with prior SSRI or TCA exposure, while SSRIs were added for those with prior TCA exposure (Supplementary Table II &III).

The prescribing profile of patients exposed to antidepressant only after stroke (Figure 2B) was broadly comparable to the ‘after’ profile for patients exposed both before and after stroke. However, we observed an increase in the frequency of mirtazapine prescriptions and a decrease in the frequency of TCA and venlafaxine prescriptions.

### 3.4. The effect of specific antidepressant classes on the functional status of patients exposed only after stroke

Disaggregation of antidepressant classes in patients exposed only after stroke showed differential effects on median ADL scores (Figure 3). Median ADL scores were highest in patients exposed to MAOIs (37.5 [IQR 31-44]) and lowest in patients exposed to TCAs (18.5 [IQR 8.25-40]) and venlafaxine (14.0 [IQR 4.25-22.75]).

**Figure 3.**
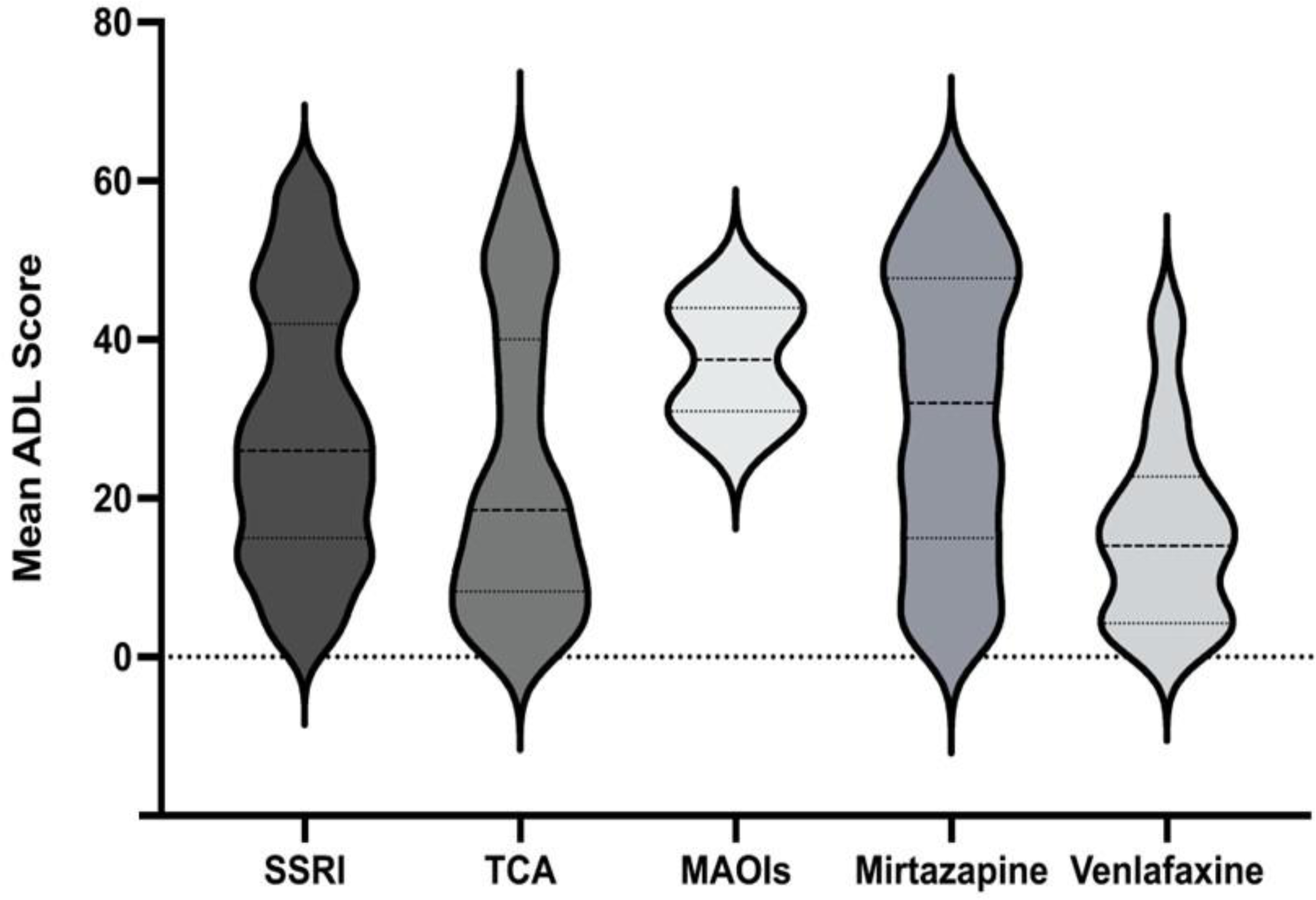
ADL score distribution in patients exposed to antidepressants after stroke varied by class. Patients on TCA and venlafaxine had lower scores, while those on mirtazapine had higher scores. Dotted lines indicate median, and solid lines represent the 1st and 3rd quartile scores. SSRI: selective serotonin reuptake inhibitor, TCA: tricyclic antidepressant, MAOI: monoamine oxidase inhibitor.

The distribution of ADL scores differed between antidepressant classes. We observed a positively skewed distribution and a higher probability of a lower ADL score in patients exposed to TCA and venlafaxine. In contrast, patients exposed to mirtazapine were more likely to have a higher ADL score.

Beta regression revealed that the ORs for TCA exposure after stroke was 0.75 (95% CI 0.59–0.96) for the unadjusted model and 0.75 (95% CI 0.58–0.95) for the adjusted model (Table 3). Both models indicate patients with TCA exposure after stroke had significantly higher odds of a lower ADL score than patients exposed to an SSRI. In addition, TCA exposure was associated with a decrease in precision and an increase in variance.

**Table 3:**
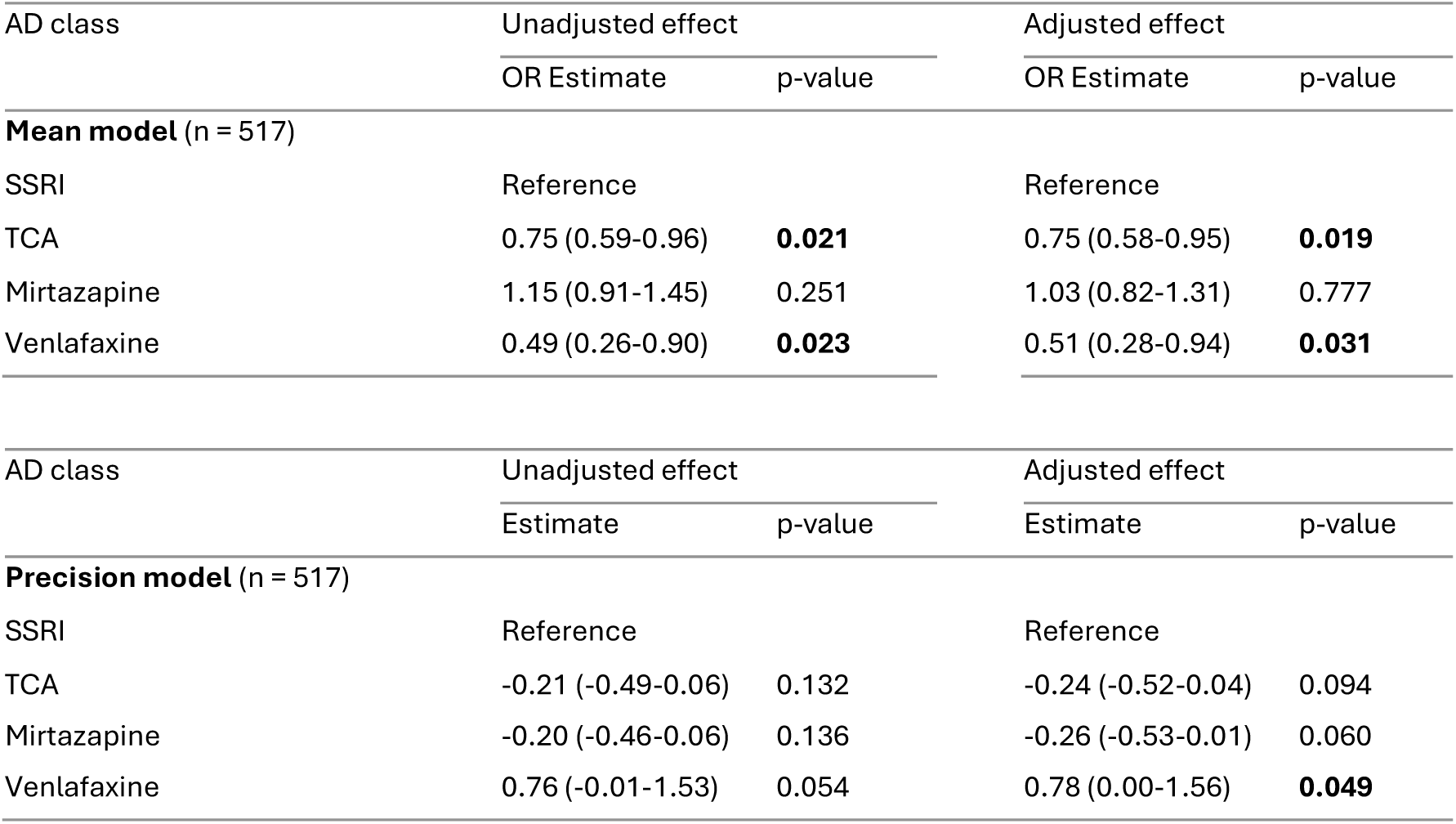
Beta regression – Antidepressant exposure only post stroke by class. Figures in parentheses are 95% confidence intervals. OR: odds ratio. The mean and precision components of the model were adjusted for age, sex, ethnicity, body mass index, length of hospital stay, and other medicines. All centrally acting medicines were considered in the mean model; the precision model used pain and antipsychotic medicines.

Similarly, the unadjusted and adjusted ORs for venlafaxine exposure were 0.49 (95% CI 0.26–0.90) and 0.51 (95% CI 0.28–0.94), respectively. For the unadjusted and adjusted models, venlafaxine exposure was associated with increased precision and decreased variance.

## 4. Discussion

The current study examined the relationship between the timing of exposure to antidepressant medicines and post-stroke functional status in a cohort of NZ patients who experienced an ischaemic stroke.

### Key clinical findings

We found that the timing of antidepressant exposure significantly affects post-stroke functional status. Patients exposed to antidepressants only after stroke had 1.3 times higher odds of poorer outcomes compared to those exposed before and after or unexposed patients. This supports the hypothesis that antidepressant exposure negatively impacts stroke outcomes. (20, 21)

The observed poorer long-term functional status in patients exposed to treatment only after stroke does not appear to be driven by a difference in prescribing patterns or patient demographics (Supplementary Table IV). Further, we observed no difference in demographics, or the length of hospital stay between exposure groups, suggesting poorer functional status was not related to greater initial stroke severity. While other factors including socioeconomic status (22) contribute to length of hospital stay, it remains a reasonable proxy for stroke severity.(15, 23).

The difference in ADL scores between patients exposed only after stroke and unexposed patients was comparable to that previously reported between patients requiring supported care and those living at home.(24) This suggests antidepressant exposure after stroke has a large enough impact on functional status to affect independence.

Post stroke depression is one of the most frequent complications after stroke. The prevalence of depression in the post-stroke period in the current study was comparable to that reported by Shi *et al* at one to five months after stroke.(25) Prescribing antidepressant medicines in a way that minimises impact on long-term functional status therefore has the potential to reduce indirectly the number of patients living with stroke-related disabilities. Our results suggest that compared to SSRI exposure, both TCAs and venlafaxine reduced the odds of poor long-term functional status. The effect of venlafaxine on ADL score was more generalisable across all individuals and less unpredictable than that of TCAs.

### Strengths and limitations

The strengths of this study include the large cohort size, which allows our results to be generalisable to the wider New Zealand population. It is also novel to use beta regression to model appropriately the skewed distribution of ADL scores and to explore the effect of specific antidepressant classes.

Our study has limitations common to observational research using health administrative data. Stroke location was unknown, and the higher incidence of left frontal lobe and basal ganglia strokes in those with post-stroke depression may lead to greater motor deficits and disability in performing ADLs. Additionally, antidepressants may be prescribed for other conditions, making it difficult to assess the cumulative impact of anxiety, insomnia, and neuropathic pain on functional status.

## Conclusion

We used population-based health administrative data to demonstrate that functional status is significantly poorer in patients who started antidepressants only after stroke compared to those who used them before and after stroke or not at all. Further, the detrimental effect of antidepressants on functional status may be mitigated in part by prescribing a TCA or venlafaxine in preference to an SSRI.

This study highlights an opportunity for healthcare professionals to improve treatment strategies that enhance both mental health and recovery outcomes for stroke survivors. Future research should focus on guiding prescribing decisions by exploring whether antidepressants have a direct effect on recovery trajectories and identifying how patient characteristics contribute to poorer long-term outcomes.

## Declarations

### Conflicts of interest/Competing interests

The authors declare no conflicts of interest.

All inferences, opinions, and conclusions drawn in this article are those of the authors, and do not reflect the opinions or policies of the Data Steward(s).

### Availability of data and material (data transparency)

Details of how data were obtained are available at the Ministry of Health (https://www.health.govt.nz/nz--statistics/national-collections-and-surveys/collections/national-minimum-dataset-hospital-events).

### Code availability (software application or custom code)

Custom code will be made available on request.

### Ethics approval

The Human Ethics Committee at the University of Otago, New Zealand reviewed and approved this study (Reference: HD24/008).

### Consent to participate

The National Ethics Advisory Committee’s Ethical Guidelines permit the re-use of de-identified Ministry of Health data for research with a written consent waiver.

### Consent for publication

The National Ethics Advisory Committee’s Ethical Guidelines permit the publication of de-identified Ministry of Health data with a written consent waiver.

### Author Contribution

ALM conceived and designed the study, supervised DG, SN and EMT and drafted and revised the paper. SN, EMT and DG provided input into the study design, performed data cleaning and analysis, drafted manuscript sections and reviewed paper revisions. RL and AJS drafted sections of the manuscript and reviewed revisions.

## Data Availability

Details of how data were obtained are available at the Ministry of Health
(https://www.health.govt.nz/nz--statistics/national-collections-and-surveys/collections/national-minimum-dataset-hospital-events).

## Acknowledgements

The authors thank Dr Bruce Russell for assistance with medicine classification.

