## Supplementary Materials for "Timing of antidepressant use influences long term functional status in New Zealand stroke patients: A retrospective population level study"

**Table I: List of medicines in AD and co-variable medication categories**

**Antidepressant medications**
Amitriptyline
Buspirone
Bupropion
Citalopram
Clomipramine
Dosulepin
Doxepin
Escitalopram
Fluoxetine
Imipramine
Maprotiline
Mianserin
Mirtazapine
Moclobemide
Nortriptyline
Paroxetine
Sertraline
Tranylcypromine
Venlafaxine

**Antiepileptic medications**
Carbamazepine
Clobazam
Clonazepam
Ethosuximide
Gabapentin
Lamotrigine
Levetiracetam
Phenobarbital
Phenytoin
Primidone
Sodium Valproate
Topiramate

**Antimuscarinic agents**
Benzatropine
Hyoscine
Procyclidine

**Antinausea medications**
Cyclizine
Metoclopramide
Metoclopramide with paracetamol
Ondansetron

**Antipsychotic medications**
Amisulpride
Aripiprazole
Chlorpromazine
Clozapine
Flupenthixol
Fluphenazine
Haloperidol
Levomepromazine
Olanzapine
Paliperidone
Pericyazine
Prochlorperazine
Quetiapine
Risperidone
Trifluoperazine
Zuclopenthixol

**Hypnotics**
Alprazolam
Diazepam
Lorazepam
Lormetazepam
Midazolam
Nitrazepam
Oxazepam
Temazepam
Triazolam
Zopiclone

**Migraine medication**
Ergotamine with caffeine
Pizotifen
Rizatriptan
Sumatriptan

**NSAID**
Aspirin
Celecoxib
Diclofenac
Ibuprofen
Ketoprofen
Naproxen
Sulindac
Tenoxicam

**Other centrally acting**
Lithium carbonate
Methylphenidate

**Paracetamol**
Paracetamol

**Pain medication**
Codeine
Dihydrocodeine
Fentanyl
Methadone
Morphine
Nefopam
Oxycodone
Paracetamol with codeine
Pethidine
Tramadol

**Smoking cessation medications**
Nicotine
Varenicline

**Parkinson's disease (PD) and Huntington's disease* (HD)**
Amantadine
Levodopa with benserazide
Levodopa with carbidopa
Pramipexole
Ropinirole
Tolcapone
Selegiline
*Tetrabenazine

**Anti-cancer drugs**
Aprepitant

**Cholinesterase inhibitors**
Donepezil
Rivastigmine

**Supplementary Results**

**Table II: Frequency of single medication switches in patients exposed both before and after stroke**

| | | Post Stroke | | |
|---|---|---|---|---|
| | | Mirtazapine | SSRI | TCA |
| Before Stroke | SSRI | 6 | 7 | 0 |
| | TCA | 3 | 7 | 6 |
| | Venlafaxine | 0 | 1 | 0 |

SSRI: selective serotonin reuptake inhibitor, TCA: tricyclic antidepressant

**Table III: Frequency of medications added in the post stroke period in patients exposed both before and after stroke**

| | | Post Stroke | | | |
|---|---|---|---|---|---|
| | | Mirtazapine | SSRI | TCA | Venlafaxine |
| Before Stroke | Mirtazapine | 0 | 1 | 1 | 1 |
| | SSRI | 8 | 2 | 5 | 2 |
| | TCA | 8 | 8 | 5 | 0 |
| | Venlafaxine | 1 | 0 | 2 | 0 |

SSRI: selective serotonin reuptake inhibitor, TCA: tricyclic antidepressant

**Table IV: Population characteristics of patients exposed only after stroke**

| **Characteristic** | **Overall** (n = 519) | **SSRI** (n = 302) | **TCA** (n = 96) | **Mirtazapine** (n = 107) | **MAOI** (n = 2) | **Venlafaxine** (n = 12) | ***p*-value** |
|---|---|---|---|---|---|---|---|
| **Age Category (Years)** | | | | | | | |
| 40-55 | 6 (1.2%) | 6 (2.0%) | 0 (0%) | 0 (0%) | 0 (0%) | 0 (0%) | |
| 56-70 | 58 (11%) | 31 (10%) | 11 (11%) | 14 (13%) | 0 (0%) | 2 (17%) | |
| 71-85 | 292 (56%) | 174 (58%) | 49 (51%) | 60 (56%) | 2 (100%) | 7 (58%) | |
| >85 | 163 (31%) | 91 (30%) | 36 (38%) | 33 (31%) | 0 (0%) | 3 (25%) | |
| **BMI Category** | | | | | | | |
| Underweight | 58 (11%) | 28 (9.3%) | 7 (7.3%) | 20 (19%) | 2 (100%) | 1 (8.3%) | |
| Normal | 136 (26%) | 70 (23%) | 29 (30%) | 34 (32%) | 0 (0%) | 3 (25%) | |
| Overweight | 236 (45%) | 146 (48%) | 44 (46%) | 39 (36%) | 0 (0%) | 7 (58%) | |
| Obese | 89 (17%) | 58 (19%) | 16 (17%) | 14 (13%) | 0 (0%) | 1 (8.3%) | |
| **Sex** | | | | | | | **0.008** |
| Male | 235 (45%) | 154 (51%) | 31 (32%) | 46 (43%) | 0 (0%) | 4 (33%) | |
| Female | 284 (55%) | 148 (49%) | 65 (68%) | 61 (57%) | 2 (100%) | 8 (67%) | |
| **Ethnic Group** | | | | | | | |
| Asian | 22 (4.2%) | 8 (2.6%) | 8 (8.3%) | 5 (4.7%) | 0 (0%) | 1 (8.3%) | |
| European | 421 (81%) | 248 (82%) | 76 (79%) | 88 (82%) | 0 (0%) | 9 (75%) | |
| Māori | 52 (10%) | 30 (9.9%) | 9 (9.4%) | 9 (8.4%) | 2 (100%) | 2 (17%) | |
| Pacific peoples | 24 (4.6%) | 16 (5.3%) | 3 (3.1%) | 5 (4.7%) | 0 (0%) | 0 (0%) | |
| **Location of Assessment** | | | | | | | 0.2 |
| HC | 464 (89%) | 268 (89%) | 90 (94%) | 94 (88%) | 1 (50%) | 11 (92%) | |
| LTCF | 55 (11%) | 34 (11%) | 6 (6.3%) | 13 (12%) | 1 (50%) | 1 (8.3%) | |
| **TLoS (days)** | 9 ± 8 | 9 ± 9 | 7 ± 6 | 10 ± 8 | 16 ± 13 | 8 ± 5 | **0.026** |
| **TTA (days)** | 148 ± 246 | 125 ± 206 | 276 ± 362 | 74 ± 94 | 66 ± 52 | 377 ± 455 | **<0.001** |

Data is presented as Mean ± SD or N (%) unless otherwise stated.
BMI body mass index, HC Home Care, LTCF Long Term Care Facility, MAOI monoamine oxidase inhibitor, SSRI selective serotonin reuptake inhibitor, TCA tricyclic antidepressant, TLoS Total Length of Hospital Stay, TTA Time to Assessment.
